# Associations between *CYP3A4, CYP3A5* and *SCN1A* Polymorphisms and Carbamazepine Metabolism in Epilepsy: A Meta-analysis

**DOI:** 10.1101/2020.03.03.20030783

**Authors:** Gui-Xin Zhao, Zheng Zhang, Wen-Ke Cai, Ming-Li Shen, Ping Wang, Gong-Hao He

**Author notes:** Correspondence to: Gong-Hao He, Ph.D., 212 Daguan Rd, Kunming 650032, China. Gui-Xin Zhao, Zheng Zhang and Wen-Ke Cai contributed equally to this work.

## Abstract

**Background and objective:** *CYP3A4* (rs2242480), *CYP3A5* (rs776746) and *SCN1A* (rs3812718 and rs2298771) gene polymorphisms were previously indicated to be associated with carbamazepine (CBZ) metabolism and resistance in epilepsy. However, previous studies regarding the effects of these polymorphisms still remain controversial. Therefore, we performed a meta-analysis to evaluate whether the four polymorphisms are associated with CBZ metabolism and resistance.

**Methods:** The PubMed, EMBASE, Cochrane library, Chinese National Knowledge Infrastructure, Chinese Science and Technique Journals Database, China Biology Medicine disc and Wan fang Database were searched up to January 2020 for appropriate studies regarding the association of rs2242480, rs776746, rs3812718 and rs2234922 polymorphisms with metabolism and resistance to CBZ. The meta-analysis was conducted by Review Manager 5.3 software.

**Results:** Eighteen studies involving 2574 related epilepsy patients were included. Significant associations between *CYP3A4* rs2242480, *CYP3A5* rs776746 and *SCN1A* rs3812718 polymorphisms and plasma concentrations of CBZ were observed. Additionally, *SCN1A* rs3812718 polymorphism was significantly associated with CBZ resistance. However, no association was observed between *SCN1A* rs2298771 polymorphism and metabolism and resistance to CBZ.

**Conclusion:** The *CYP3A4* rs2242480, *CYP3A5* rs776746 and *SCN1A* rs3812718 polymorphisms may play important roles in metabolism and resistance to CBZ, while *SCN1A* rs2298771 polymorphism is not associated with CBZ in epilepsy. These findings would improve the individualized therapy of epileptic patients in clinics.

## 1. INTRODUCTION

Epilepsy, one of the most common chronic neurological disorder, affects more than 70 million people worldwide[1]. In addition, long-term recurrent seizures was associated with increased morbidity and mortality, which can give rise to a large social and economic burden and seriously hurt patients’ physical and mental health[2]. For most people with epilepsy, anti-epileptic drug (AED) monotherapy is a preferred treatment modality, with the aim of reducing risks for drug–drug interactions, fewer adverse events, and better patient compliance[3, 4].

Carbamazepine (CBZ) therapy is one of the most economical treatment usually prescribed, exhibiting beneficial efficacy in control of simple partial, complex partial, and generalized tonic-clonic seizures and mitigating long-term mortality[5]. However, from the pharmacological point of view, CBZ is a powerful liver enzyme inducer, with the characteristic of inducing self-metabolism and hence resulting in the individualized differences of clinical efficacy[6]. Moreover, the metabolism of CBZ is a complex oxidative and epoxidase pathway. First, CBZ is mainly catalyzed by cytochrome P450 (CYP) 3A4 and 3A5 to produce active metabolite carbamazepine-10,11-epoxide (CBZE), which possesses a potent anticonvulsant effect[7]. Subsequently, CBZE is hydrolyzed via microsomal epoxide hydrolase, which is encoded by the *EPHX1* gene, to an inactive metabolite carbamazepine-10,11-trans dihydrodiol (CBZD)[7-9]. It was previously reported that genetic variation of drug-metabolizing enzymes could generate the inter-individual differences in the pharmacokinetics and pharmacodynamics of epileptic patients[10-12]. In our recent meta-analysis, it was further proved that *EPHX1* polymorphisms (rs1051740 and rs2234922) were significantly associated with concentrations of both CBZ and CBZE and metabolism ratio[13]. However, previous studies regarding the effects of polymorphisms of *CYP3A4/3A5* on the pharmacokinetics and pharmacodynamics of CBZ still remain controversial. For example, it was shown that *CYP3A4*\*1G (g.20230G>A, rs2242480) variant carriers had lower plasma concentration of CBZ than *CYP3A4*\*1/*1 carriers[14, 15]. On the contrary, several other studies found no significant associations between the variant *CYP3A4* and plasma concentration of CBZ[16, 17]. Additionally, similar inconsistent results were also found for polymorphism of *CYP3A5* (g.6986A>G, rs776746). One study reported that the *CYP3A5* rs776746 polymorphism had no associations with plasma concentration of CBZ[18], but several other studies revealed that *CYP3A5* rs776746 nonexpressers had higher plasma concentration of CBZ than expressers[19, 20].

In addition, CBZ is a sodium channel blocker and mainly inhibit voltage-sensitive sodium channel activity[21]. Its mechanism of action was binding to a receptor site in the pore of neuronal sodium channels, decreasing sodium influx, reducing the number and duration of action potentials within a burst and thereby reducing the excitability of the central neurons[22, 23]. The voltage-gated sodium channel α subunit type 1 (SCN1A) is a heterologous complex, which comprises a large hollow glycosylation α subunit encoded by *SCN1A* gene and two small affiliated β subunits encoded by *SCN2A* gene[24, 25]. Previous studies indicated that variations in sensitivity of pharmacological targets might arise from single nucleotide polymorphisms (SNPs)[26-28]. However, these reports did not clarify the correlations between the *SCN1A* rs3812718 (IVS5 G>A) and rs2298771 (c.3184A>G) polymorphisms and resistance to CBZ in epilepsy. For example, it was shown that the rs3812718 and rs2298771polymorphisms were significantly associated with CBZ resistance in English and Chinese cohorts of patients[29, 30]. On the contrary, one study found that the variant *SCN1A* had no significant influence on CBZ resistance in Japanese epilepsy patients[31].

Therefore, it is necessary to comprehensively evaluate the associations of the abovementioned polymorphisms in *CYP3A4, CYP3A5* and *SCN1A* genes with CBZ metabolism and resistance to achieve a more rational and individualized use of CBZ in epileptic treatment. In this regard, a systematic meta-analysis based on all currently eligible studies was hence performed in this study hoping to provide novel insights into the roles of genetic variations on the pharmacokinetics and pharmacodynamics of CBZ.

## 2. MATERIALS AND METHODS

### 2.1. Search strategy

The recommendations of the Preferred Reporting Items for Systematic reviews and Meta-Analyses (PRISMA) statement were followed in this study[32]. A comprehensive literature search of studies in all languages up to January 2020 across PubMed, EMBASE, Cochrane library, Chinese National Knowledge Infrastructure (CNKI), Chinese Science and Technique Journals Database (VIP), China Biology Medicine disc (CBM) and Wan fang Data Information Site was conducted.

Relevant studies were identified using the terms: “carbamazepine”, “CYP 450” or “Cytochrome P-450 Enzyme System”, “SCN1A protein” or “NAV1.1 Voltage-Gated Sodium Channel”, “polymorphism” or “genotype” and “epilepsy”. The search strategies for PubMed were provided in supplementary materials. In addition, we carried out a manual retrieve of references of all acquired articles to identify additional eligible studies. The literature retrieval was performed by two independent authors and the discrepancy was resolved by consensus.

### 2.2. Inclusion and exclusion criteria

Studies fulfilling the following criteria were considered as eligible for inclusion in this meta-analysis: (a) original studies that investigated the associations between the polymorphisms of *SCN1A, CYP3A4* or *CYP3A5* and plasma concentration or resistance of CBZ in epilepsy, (b) each patient was administrated with CBZ monotherapy, (c) no liver or renal diseases were reported, and (d) including at least three studies for each polymorphism to allow calculation of publication bias between studies. Exclusion criteria were the following: (a) insufficient information for data extraction, (b) reviews, case reports or articles only with an abstract, and (c) studies in vitro experiments.

### 2.3. Data extraction

Two reviewers independently extracted data included authors, publication date, country, sample size, allelic and genotype distribution, plasma concentration of CBZ, CBZE and CBZD, CBZE:CBZ, CBZD:CBZ and CBZD:CBZE ratios, drug resistant and responsive patients. According to the criteria International League Against Epilepsy (ILAE) 2010, drug resistant epilepsy was defined as failure of adequate trials of two tolerated and appropriately chosen and used AED schedules (whether as monotherapies or in combination) to achieve sustained seizure freedom; drug-responsive epilepsy was defined as epilepsy in which the patients receiving the current AED treatment regimen has been seizure free for a minimum of three times the longest pre-intervention interseizure interval or 12 months, whichever is longer[33]. The authors of the studies were contacted for additional data when necessary and applicable. Disagreement between the reviewers was resolved by discussion or consensus with the third investigator.

### 2.4. Assessment of methodological quality

The Newcastle-Ottawa Scale (NOS), which was developed to assess the quality of non-randomized studies in the meta-analysis, was used to assess the methodological quality of included studies[34]. The NOS used a star rating system to judge quality based on eight items and categorized into three broad perspectives: selection, comparability and exposure. High-quality studies were identified with a NOS score of five or more, whereas those with less than a score of five were considered as low-quality studies. The quality assessment was performed by two investigators independently, and disagreements were resolved by discussion and consensus.

### 2.5. Statistical analysis

The Review Manager 5.3 software was utilized for statistical analysis. Comparisons of the dichotomous variables were performed using risk ratio (RR) with 95% confidence interval (CI). For continuous variables, the standard mean differences (SMD) or mean differences (MD) with 95% CI were used in the meta-analysis. The statistical significance was determined by *Z* test, and *P*-value < 0.05 was considered as statistically significant. Heterogeneity was tested using the Cochran’s *Q* test and measured inconsistency by *I*^*2*^[35]. Data with low heterogeneity (*P* > 0.10 and *I*^*2*^ < 50%) were analyzed by a fixed-effects model while a random-effect model was used for data with high heterogeneity (*P* ≤ 0.10 and *I*^*2*^ ≥ 50%). Sensitivity analyses were performed by excluding one study at a time to explore sources of heterogeneity and evaluate the stability of pooled results. Moreover, a funnel plot was conducted to assess the publication bias when applicable.

## 3. RESULTS

### 3.1. Study selection and qualitative assessment

Detailed flow diagram of the study selection process is presented in Fig. 1. The initial search strategy identified 819 studies in the electronic databases, from which 53 were excluded for duplication. Then, we excluded 710 studies after reading titles, abstracts and full-text articles of the remaining 766 studies, which included 459 irrelevant studies, 251 review articles or commentaries, 13 in vitro or animal studies, 25 studies with insufficient data. Eventually, 18 studies were included according to the present inclusion and exclusion criteria. Of the included studies, the polymorphisms of rs2242480, rs776746, rs3812718 and rs2298771 were reported in 4[14-17], 10[14-16, 18, 20, 36-40], 7[16, 17, 30, 41-44] and 6[16, 30, 41, 42, 45, 46] studies, respectively.

**Fig. 1.**
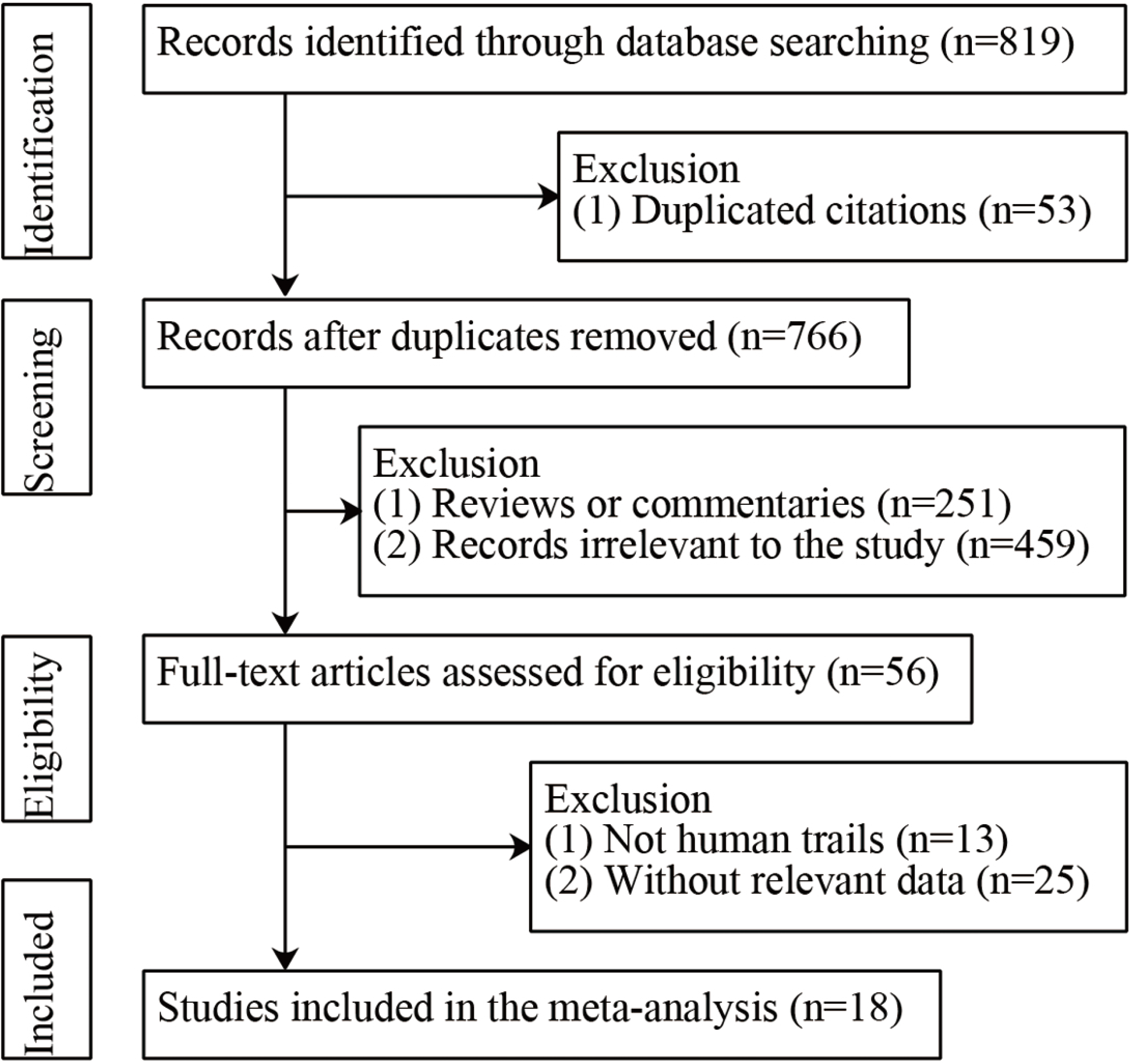
Flow diagram for process of included studies selection.

A total of 2574 patients were included in this meta-analysis. The characteristics from selected studies in *CYP3A4, CYP3A5* and *SCN1A* polymorphisms and CBZ metabolism and resistance were summarized in Table 1. These studies were published between 2009 and 2019 from different countries. All participants in the studies were epilepsy patients and were treated with CBZ monotherapy at a stable maintenance dose. The scores ranging from 5 to 8 based on the NOS evaluation system, indicating a relatively high quality of included studies. Furthermore, the publication bias was not conducted due to the insufficient number of studies in the present study[47].

**Table 1.** Characteristics of all studies included in this meta-analysis.

| First author | Year | Population | Subjects | Drug | Treatment duration | Number<br>(male/female) | Age/years<br>(M ± SD) | Weight/Kg<br>(M ± SD) | NOS score | Gene | Polymorphism | Ref. |
| --- | --- | --- | --- | --- | --- | --- | --- | --- | --- | --- | --- | --- |
| Ma CL | 2015 | China | Epilepsy | CBZ monotherapy | Over 1 year | 166(91/75) | 34.75±15.83 | 64.96±11.31 | 8 | CYP3A4<br>CYP3A5<br>SCN1A | rs2242480<br>rs776746<br>rs3812718<br>rs2298771 | [16] |
| Park PW | 2009 | Korea | Epilepsy | CBZ monotherapy | Over 3 months | 35(27/8) | 34.66±13.23 | 65.44 ± 14.93 | 5 | CYP3A5 | rs776746 | [20] |
| Meng HM | 2011 | China | Epilepsy | CBZ monotherapy | Over 1 week | 84(53/31) | 36.11±16.47 | NR | 7 | CYP3A5 | rs776746 | [36] |
| Yun W | 2013 | China | Epilepsy | CBZ monotherapy | Over 1 year | 83(53/30) | 36.2±16.5 | 63.29±7.65 | 8 | CYP3A4<br>SCN1A | rs2242480<br>rs3812718 | [17] |
| Zhu X | 2014 | China | Epilepsy | CBZ monotherapy | Over 1 year | 210(119/91) | 19±15.09 | 49.22±16.6 | 8 | CYP3A4<br>CYP3A5 | rs2242480<br>rs776746 | [14] |
| Liu JF | 2014 | China | Epilepsy | CBZ monotherapy | Over 3 months | 40(24/16) | 41.08±17.91 | 59.67±8.97 | 5 | CYP3A5 | rs776746 | [37] |
| Liu X | 2016 | China | Epilepsy | CBZ monotherapy | Over 3 months | 67(46/21) | 11.5±4.63 | 44.12±16.29 | 5 | CYP3A5 | rs776746 | [18] |
| Wang JH | 2012 | China | Epilepsy | CBZ monotherapy | Over 2 months | 177(100/77) | 31.41±13.38 | 61.01±8.39 | 7 | CYP3A4<br>CYP3A5 | rs2242480<br>rs776746 | [15] |
| Wang P | 2014 | China | Epilepsy | CBZ monotherapy | Over 1 month | 66(37/29) | 33.03±13.52 | 60.03±12.59 | 5 | CYP3A5 | rs776746 | [38] |
| Zhang Q | 2016 | China | Epilepsy | CBZ monotherapy | Over 1 month | 278(148/130) | 38.19±16.96 | 66.04±8.85 | 6 | CYP3A5 | rs776746 | [39] |
| Zhu YM | 2012 | China | Epilepsy | CBZ monotherapy | Over 1 week | 58(36/22) | NR | NR | 5 | CYP3A5 | rs776746 | [40] |
| Daci A | 2015 | Kosovo | Epilepsy | CBZ monotherapy | Over 6 months | 145(82/63) | 32.9±15.5 | 68.3±16 | 6 | SCN1A | rs3812718<br>rs2298771 | [41] |
| Hung CC | 2012 | China | Epilepsy | CBZ monotherapy | Over 7 months | 234(111/123) | 39.23±11.88 | 60.12±10.51 | 6 | SCN1A | rs3812718<br>rs2298771 | [42] |
| Nazish HR | 2018 | Pakistan | Epilepsy | CBZ monotherapy | Over 6 months | 80(39/41) | 18.7±9.5 | NR | 7 | SCN1A | rs2298771 | [45] |
| Sterjev Z | 2012 | Macedonia | Epilepsy | CBZ monotherapy | Over 1 year | 147(63/84) | NR | NR | 6 | SCN1A | rs3812718 | [43] |
| Namazi S | 2015 | Iran | Epilepsy | CBZ monotherapy | Over 1 year | 70(27/43) | NR | 57±11.3 | 5 | SCN1A | rs2298771 | [46] |
| Huang JM | 2019 | China | Epilepsy | CBZ monotherapy | Over 1 year | 186 | NR | NR | 6 | SCN1A | rs3812718 | [44] |
| Zhou BT | 2012 | China | Epilepsy | CBZ monotherapy | Over 1 year | 448(229/219) | 30.4±16.7 | 49.3±9.5 | 8 | SCN1A | rs3812718<br>rs2298771 | [30] |

NOS Newcastle–Ottawa scale, $M \pm SD$ mean ± standard deviation, Ref. reference, CBZ carbamazepine, NR not reported.

### 3.2. Meta-analysis of the associations between CYP3A4 rs2242480 polymorphism and CBZ metabolism

The results of the associations between *CYP3A4* rs2242480, *CYP3A5* rs776746, *SCN1A* rs3812718 and rs2298771 polymorphisms and CBZ metabolism for meta-analysis were showed in supplemental Table S.1. Meta-analysis of the *CYP3A4* rs2242480 polymorphism showed significant associations with plasma concentration of CBZ in the heterozygote codominant model (AG vs. GG, SMD = −0.27, 95% CI = −0.43 to −0.11, *P* = 0.001) (Fig. 2c), dominant model (AA+AG vs. GG, SMD = −0.23, 95% CI = −0.39 to −0.08, *P* = 0.003) (Fig. 2d) and overdominant model (AG vs. AA+GG, SMD = −0.28, 95% CI = −0.44 to −0.12, *P* = 0.0006) (Fig. 2f), indicating that this SNP might decrease the plasma CBZ concentration in epilepsy. Moreover, one study showed that in homozygote codominant model and recessive model the ratio of CBZE to CBZ was significantly lower in carriers of the AA genotype than carriers of the GG genotype (AA vs. GG, MD = −0.04, 95% CI = −0.08 to −0.00, *P* = 0.04) (supplemental Fig. S.1b) and AG+GG genotype (AA vs. AG+GG, MD = −0.04, 95% CI = −0.08 to −0.00, *P* =0.03) (supplemental Fig. S.1e)[16]. Regarding the plasma concentrations of CBZE and CBZD, no association was found in all the genetic models (supplemental Fig. S.2 and S.3).

**Fig. 2.**
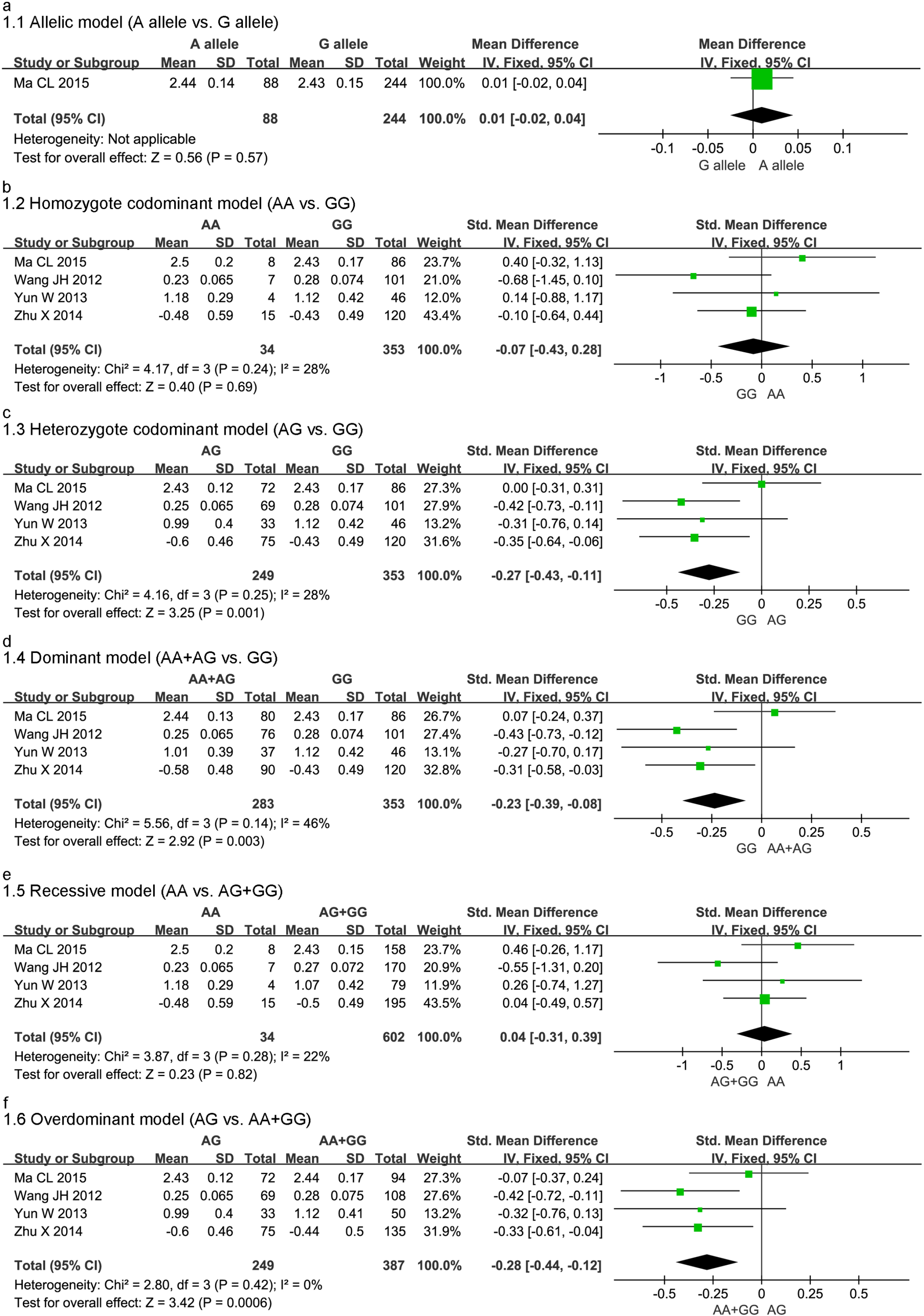
Forest plot for association between CYP3A4 rs2242480 polymorphism and plasma CBZ concentration in allelic model (a), homozygote codominant model (b), heterozygote codominant model (c), dominant model (d), recessive model (e) and overdominant model (f).

### 3.3. Meta-analysis of the associations between CYP3A5 rs776746 polymorphism and CBZ metabolism

For rs776746 polymorphism, the GG genotype and GG+GA genotype was found to be significantly associated with plasma concentration of CBZ in homozygote codominant model (GG vs. AA, SMD = 0.68, 95% CI = 0.37 to 1.00, *P* < 0.0001) (Fig. 3b), dominant model (GG+GA vs. AA, SMD = 0.38, 95% CI = 0.05 to 0.72, *P* = 0.03) (Fig. 3d) and recessive model (GG vs. GA+AA, SMD = 0.57, 95% CI = 0.21 to 0.92, *P* = 0.002) (Fig. 3e). In addition, the sensitivity analysis was also conducted by excluding each study successively, which abrogated the heterogeneity but did not influence the present results (Fig. 3g and Fig 3h). As for the plasma concentrations of CBZE, CBZD and the ratio of CBZD to CBZ, our meta-analysis did not observed any associations between rs776746 polymorphism and the metabolism of CBZ in all the genetic models (supplemental Fig. S.4-S.6). However, when sensitivity analysis for recessive model of the plasma concentrations of CBZE was performed, the association became significant with the heterogeneity relatively reduced (SMD = −0.49, 95% CI = −0.82 to −0.17, *P* = 0.003) (supplemental Fig. S.4g).

**Fig. 3.**
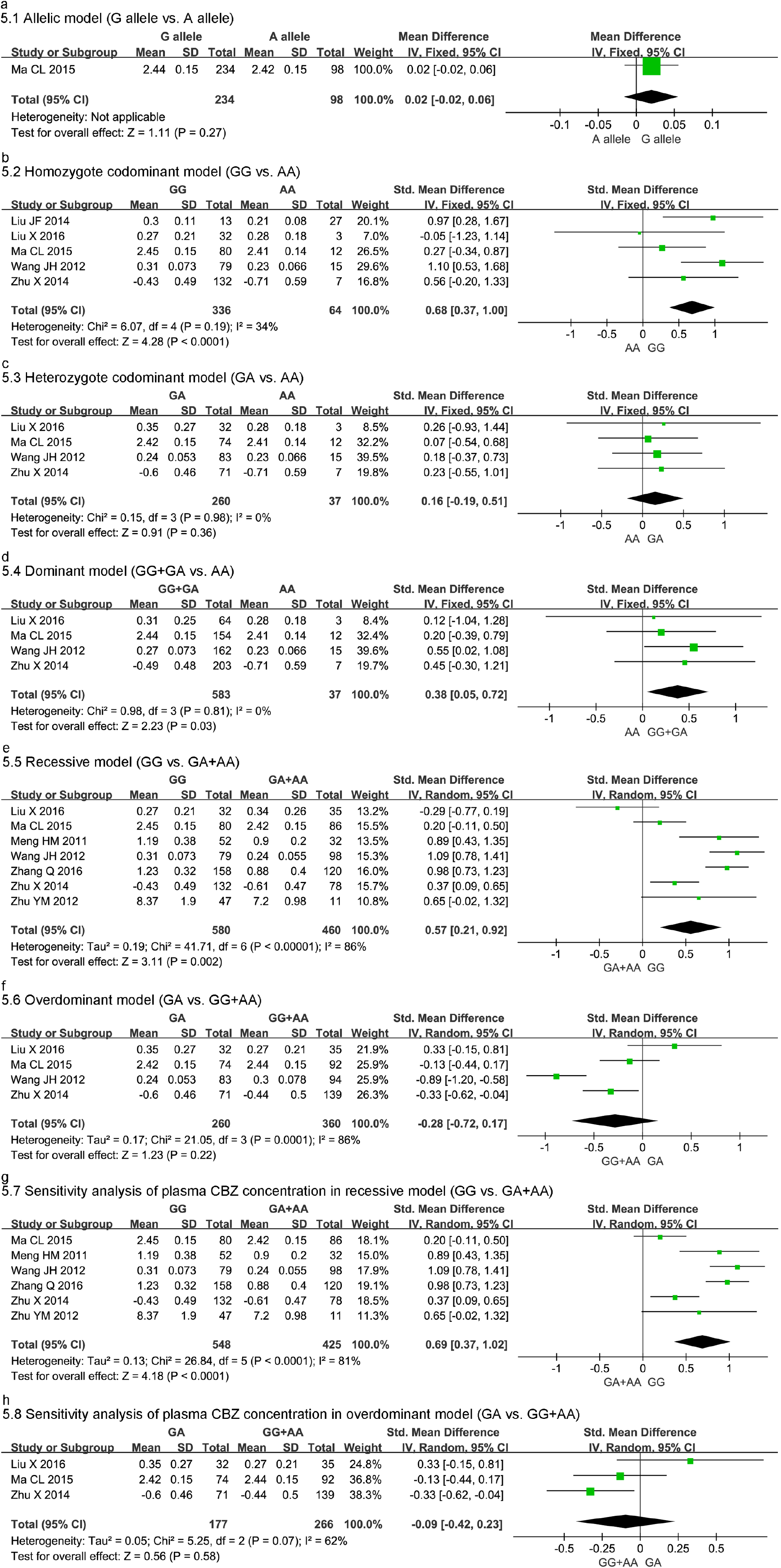
Forest plot for association between CYP3A5 rs776746 polymorphism and plasma CBZ concentration in allelic model (a), homozygote codominant model (b), heterozygote codominant model (c), dominant model (d), recessive model (e) and overdominant model (f). Sensitivity analysis for effect of CYP3A5 rs776746 polymorphism on plasma CBZ concentration in recessive model (g) and overdominant model (h).

### 3.4. Meta-analysis of the associations between SCN1A rs3812718 and rs2298771 polymorphisms and CBZ metabolism

For rs3812718, the allelic model (A allele vs. G allele, SMD = −0.58, 95% CI = −0.73 to −0.44, *P* < 0.00001) (Fig. 4a) and the overdominant model (AG vs. AA+GG, SMD = −0.19, 95% CI = −0.30 to −0.08, *P* = 0.001) (Fig. 4f) showed a significant difference between rs3812718 polymorphism and plasma concentration of CBZ. The sensitivity analysis decreased the heterogeneity but did not influence the present results (supplemental Fig. S.7). In addition, a significant difference was found in the ratio of CBZE to CBZ in heterozygote codominant model (AG vs. GG, OR = 0.32, 95% CI = 0.05 to 0.59, *P* = 0.02) (supplemental Fig. S.8c) and dominant model (AA+AG vs. GG, OR = 0.29, 95% CI = 0.03 to 0.55, *P* = 0.03) (supplemental Fig. S.8d). Furthermore, one study reported that AA genotype increased the ratio of CBZD to CBZ in homozygote codominant model (AA vs. GG, MD = 0.20, 95% CI = 0.11 to 0.29, *P* < 0.0001) (supplemental Fig. S.9a), dominant model (AA+AG vs. GG, MD = 0.07, 95% CI = 0.01 to 0.13, *P* = 0.02) (supplemental Fig. S.9c) and recessive model (AA vs. AG+GG, OR = 0.18, 95% CI = 0.09 to 0.27, *P* < 0.0001) (supplemental Fig. S.9d)[41]. As for the plasma concentrations of CBZE and CBZD and the ratio of CBZD to CBZE, no statistically significant association was found between SCN1A rs3812718 polymorphism and CBZ metabolism (supplemental Fig. S.10-S.12). For rs2298771, no statistically significant association was found between this polymorphism and CBZ metabolism in all the six genetic models (supplemental Fig. S.13-S.19). Furthermore, sensitivity analysis decreased the heterogeneity but did not alter the initial result (supplemental Fig. S.13g and S.15).

**Fig. 4.**
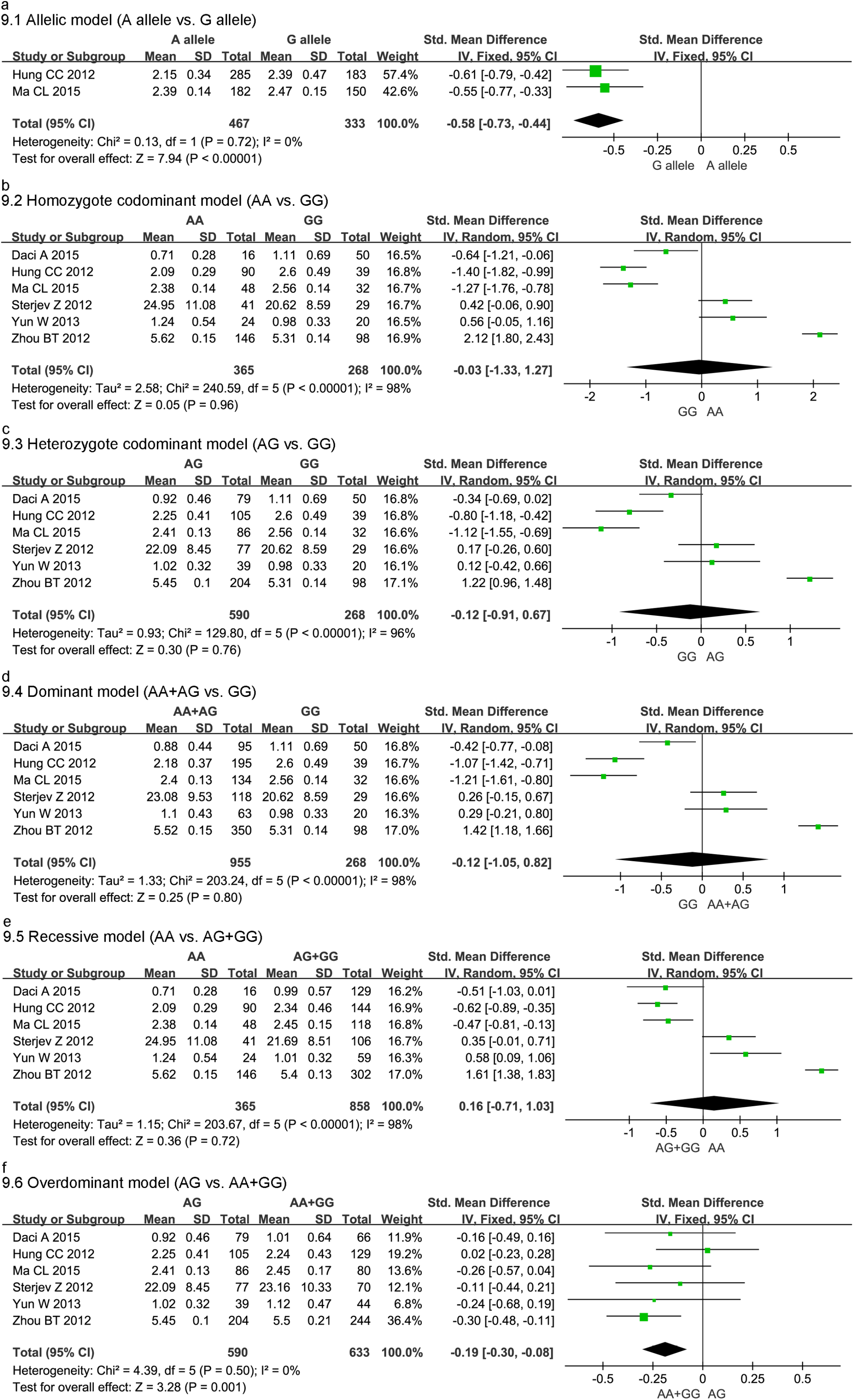
Forest plot for association between SCN1A rs3812718 polymorphism and plasma CBZ concentration in allelic model (a), homozygote codominant model (b), heterozygote codominant model (c), dominant model (d), recessive model (e) and overdominant model (f).

### 3.5. Meta-analysis of the associations between CYP3A4 rs2242480, CYP3A5 rs776746, SCN1A rs3812718 and rs2298771 polymorphisms and CBZ resistance

The association between rs2242480, rs776746, rs3812718 and rs2298771 polymorphisms and CBZ resistance was pooled in supplemental Table S.2. For *SCN1A* rs3812718, the results showed that the A allele and mutant-type homozygote AA genotype increased CBZ resistance in allelic model (A allele vs. G allele, RR = 1.18, 95% CI = 1.05 to 1.33, *P* = 0.007) (Fig 5a), homozygote codominant model (AA vs. GG, RR = 1.24, 95% CI = 1.08 to 1.43, *P* = 0.003) (Fig. 5b) and recessive model (AA vs. AG+GG, RR = 1.47, 95% CI = 1.22 to 1.77, *P* < 0.0001) (Fig. 5e). However, we did not find any association regarding *CYP3A4* rs2242480, *CYP3A5* rs776746 and *SCN1A* rs2298771 polymorphisms (supplemental Fig. S.20–S.22), indicating that these polymorphisms might not the causal factors associated with resistance of CBZ in epilepsy. Furthermore, the sensitivity analysis also effectively abrogated the heterogeneity but did not influence the present results (supplemental Fig. S22g).

**Fig. 5.**
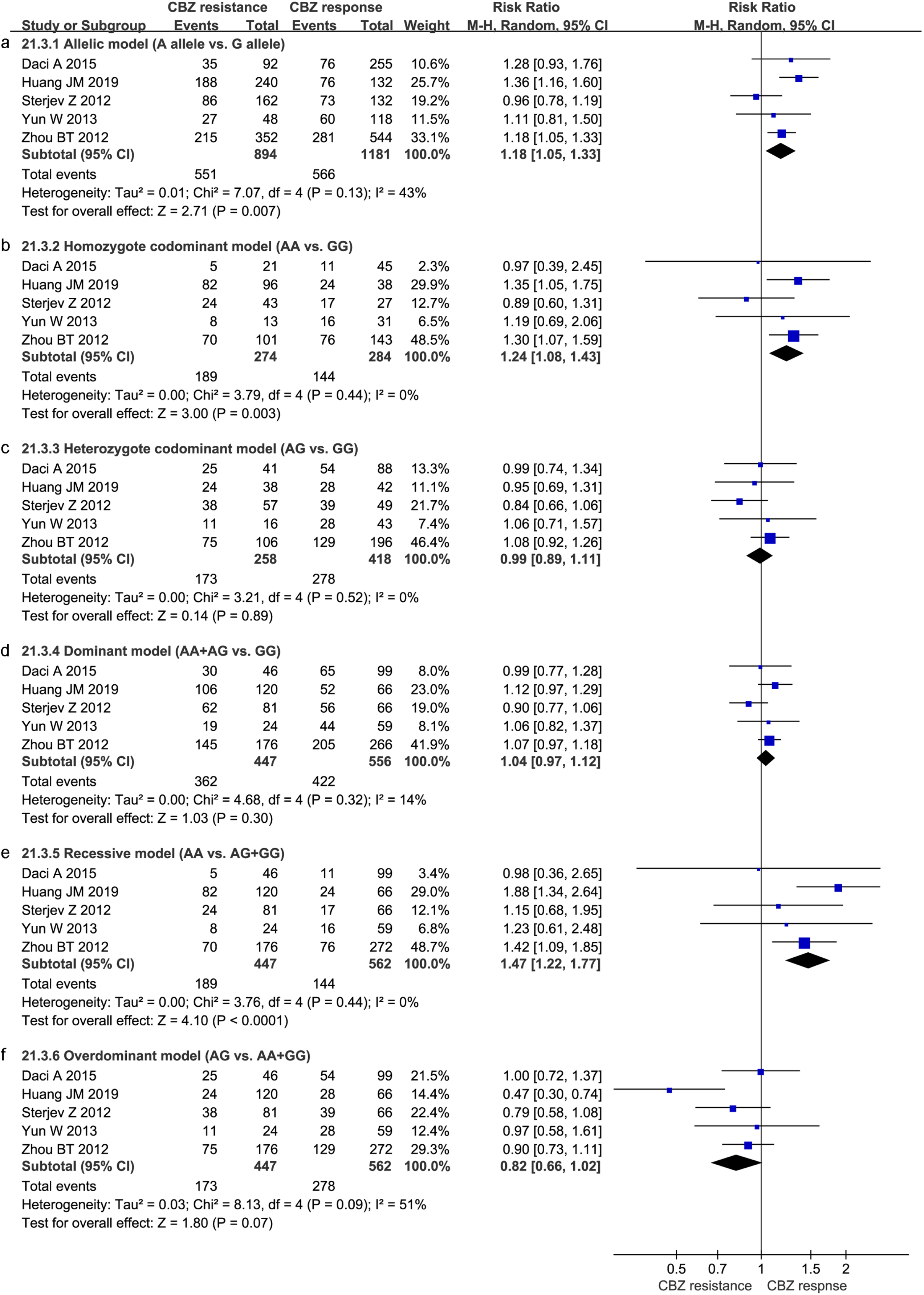
Forest plot for association between SCN1A rs3812718 polymorphism and CBZ resistance in allelic model (a), homozygote codominant model (b), heterozygote codominant model (c), dominant model (d), recessive model (e) and overdominant model (f).

## 4. DISCUSSION

With the development of pharmacogenomics, it has been shown that the variability in individual response to antiepileptic drugs depends not only on non-genetic factors (i.e., age, environment and concomitant medications), but also on genetic factors[48]. In a previous study, it was found that genetic factors accounted for 20 to 95 percent of the factors of individual differences in drug efficacy[49]. Hence, to provide a comprehensive profile of the associations of certain genetic polymorphisms with CBZ pharmacokinetics and pharmacodynamics would contribute to a more understanding of pharmacological characters of CBZ. To our knowledge, this is the first meta-analysis to address the associations between *CYP3A4, CYP3A5* and *SCN1A* polymorphisms and metabolism and efficacy of CBZ and found significant relationships between rs2298771, rs2242480 and rs776746 polymorphisms and plasma concentration of CBZ, which might further explain individual differences of CBZ and provide effective references for personalized CBZ therapy in epileptic patients.

The *SCN1A* gene encodes the alpha subunit Nav1.1 of the voltage-gated sodium channel and plays a crucial role in the pharmacological effects of CBZ[50, 51]. This gene is located on chromosome 2q24.3, consists of 26 exons and spans 91480bp of DNA[52, 53]. Alternative splicing of Nav genes has been shown to generate channels with unique functional and pharmacologic properties[54]. Previous pharmacogenomics studies demonstrated that rs3812718 genetic variant was located within an intron splice donor site and altered the proportion of *SCN1A* transcripts incorporating the canonical (exon 5A) or alternative (exon 5N) exon 5[55, 56]. Previous initial reports suggested that exon 5N was expressed primarily during the neonatal period, whereas the exon 5A was the adult exon[56-58]. Further studies revealed that the G allele of rs3812718 allowed both the 5N and 5A forms of this exon to be alternatively expressed, while the A allele reduced expression of the 5N form relative to the 5A form[55, 59, 60]. In this meta-analysis, we found that the rs3812718 SNP was significantly associated with the plasma concentration of CBZ, CBZE:CBZ ratio, CBZD:CBZ ratio and CBZ resistance, which provided further evidence that this SNP influenced the relative expression of neonatal and adult transcripts of *SCN1A*. The rs3812718 polymorphism may serve as a biomarker for individualized CBZ therapy. However, the exact underlying mechanisms of how this SNP affects the potential associations remain unclear and further functional investigation is needed to clarify this issue.

The nonsynonymous mutation rs2298771 within exon 16 of the *SCN1A* gene is characterized by substitution of alanine for threonine at the amino acid position 1067 (Thr1067Ala) and is an A to G SNP [61]. In the present meta-analysis, we failed to detect any statistically significant association of the genetic variant with CBZ metabolism and resistance, which was contrary to certain previous findings[24, 38, 62]. Several studies found that this polymorphism might affect structure-function relationship of the channel leading to misfiring of brain neurons and affecting drug efficacy and patient response[62, 63]. The inconsistencies between the present meta-analysis data and these fundamental studies may be due to the effects of various other confounded factors, including ethnicity, age, environment of the patients and physicians’ bias as well[12, 64]. Regretfully, we did not perform further subgroup analyses of these possible underlying factors because of data insufficiency. Additionally, the limited sample size might also have underestimated subtle effects of the genetic variant. Therefore, further well-designed investigations are required to verify these findings.

The cytochrome P450 gene is one of the most extensively studied genes with pharmacogenomics relevance due to its involvement in the metabolism of conventional drugs, most of which have narrow therapeutic indexes[65, 66]. Meanwhile, *CYP3A4* and *CYP3A5* were considered to play a major role during the hepatic oxidation process of CBZ[7, 67]. Among the variants within *CYP3A4* and *CYP3A5* genes, the rs2242480 polymorphism, also known as *CYP3A4*\*1G, is located in intron 10 of *CYP3A4* while the rs776746 polymorphism, also known as *CYP3A5*\*3, is located in intron 3 of *CYP3A5*[68-70]. Previous studies reported that these 2 genetic variations were respectively associated with the *CYP3A4* or *CYP3A5* enzyme activities in epileptic patients. As a functional locus, the rs2242480 polymorphism was demonstrated to enhance the expression and enzyme activity of *CYP3A4* in human liver and blood brain barrier and hence to affect the metabolism and resistance of CBZ[71, 72]. With regard to the rs776746, it was found to play a major role in the expression of the *CYP3A5* enzyme via a splicing defect of mRNA and producing a truncated and nonfunctional protein[20, 73]. In this mate-analysis, the rs2242480 polymorphism was observed to be associated with decreased serum concentration of CBZ and decreased CBZE:CBZ ratio, while the rs776746 polymorphism was observed to be associated with increased serum concentration of CBZ and decreased serum concentration of CBZE. The findings were consistent with the previous studies and further suggested that these 2 SNPs respectively affected the metabolism of CBZ[16, 55].

Some limitations of the present meta-analysis deserve consideration. Firstly, included studies recruited relatively few participants and thus further confirmatory evidence from large sample trials is required. Secondly, publication bias might not be avoided for only including published articles while missing certain related unpublished studies with negative results. Thirdly, CBZ metabolism and pharmacoresistance might be influenced by several confounding factors such as age, sex, ethnicity and interactions between SNPs. But original data deficiency limited our further assessment of these factors. Finally, there is fairly significant heterogeneity in partial outcomes of the study, which might reduce the validity of the results.

In conclusion, our meta-analysis indicated that the *SCN1A* rs3812718, *CYP3A4* rs2242480 and *CYP3A5* rs776746 polymorphisms were related to the metabolism of CBZ, while *SCN1A* rs2298771 had no obvious effects on CBZ metabolism for epilepsy. We also found that the *SCN1A* rs3812718 polymorphism was associated with an increased resistance of CBZ. These findings confirmed significant effects of *SCN1A, CYP3A4* and *CYP3A5* gene polymorphisms on pharmacokinetics and pharmacodynamics of CBZ and would improve the individualized therapy of epileptic patients in clinics.

## Data Availability

The data used to support the findings of this study are included within the article.

## ETHICS APPROVAL AND CONSENT TO PARTICIPATE

All results and analyses were based on previous ethically-approved studies, thus no further ethical approval and patient consent are required.

## HUMAN AND ANIMAL RIGHTS

This article does not contain any studies with human or animal subjects performed by any of the authors.

## ACKNOWLEDGEMENTS

Declared none.

## SUPPLEMENTARY MATERIAL

Supplementary material is available on the publisher’s website along with the published article.

## References

1. Thijs RD, Surges R, O’Brien TJ, Sander JW. Epilepsy in adults. Lancet (London, England). 2019;393(10172):689–701.

2. Strzelczyk A, Griebel C, Lux W, Rosenow F, Reese J-P. The Burden of Severely Drug-Refractory Epilepsy: A Comparative Longitudinal Evaluation of Mortality, Morbidity, Resource Use, and Cost Using German Health Insurance Data. Frontiers in neurology. 2017;8:712.

3. Shorvon SD. Drug treatment of epilepsy in the century of the ILAE: the second 50 years, 1959-2009. Epilepsia. 2009;50 Suppl 3:93–130.

4. St Louis EK, Rosenfeld WE, Bramley T. Antiepileptic drug monotherapy: the initial approach in epilepsy management. Current neuropharmacology. 2009;7(2):77–82.

5. Glauser T, Ben-Menachem E, Bourgeois B, et al. Updated ILAE evidence review of antiepileptic drug efficacy and effectiveness as initial monotherapy for epileptic seizures and syndromes. Epilepsia. 2013;54(3):551–63.

6. Perkin GD. Trigeminal Neuralgia. Current treatment options in neurology. 1999;1(5):458–65.

7. Kerr BM, Thummel KE, Wurden CJ, et al. Human liver carbamazepine metabolism. Role of CYP3A4 and CYP2C8 in 10,11-epoxide formation. Biochem Pharmacol. 1994;47(11):1969–79.

8. Thorn CF, Leckband SG, Kelsoe J, et al. PharmGKB summary: carbamazepine pathway. Pharmacogenet Genomics. 2011;21(12):906–10.

9. Makmor-Bakry M, Sills GJ, Hitiris N, et al. Genetic variants in microsomal epoxide hydrolase influence carbamazepine dosing. Clin Neuropharmacol. 2009;32(4):205–12.

10. Grover S, Gourie-Devi M, Baghel R, et al. Genetic profile of patients with epilepsy on first-line antiepileptic drugs and potential directions for personalized treatment. Pharmacogenomics. 2010;11(7):927–41.

11. Ferraro TN, Buono RJ. The relationship between the pharmacology of antiepileptic drugs and human gene variation: an overview. Epilepsy & behavior : E&B. 2005;7(1):18–36.

12. Tomalik-Scharte D, Lazar A, Fuhr U, Kirchheiner J. The clinical role of genetic polymorphisms in drug-metabolizing enzymes. The pharmacogenomics journal. 2008;8(1):4–15.

13. Zhao GX, Shen ML, Zhang Z, et al. Association between EPHX1 polymorphisms and carbamazepine metabolism in epilepsy: a meta-analysis. International journal of clinical pharmacy. 2019;41(6):1414–28.

14. Zhu X, Yun WT, Sun XF, et al. Effects of major transporter and metabolizing enzyme gene polymorphisms on carbamazepine metabolism in Chinese patients with epilepsy. Pharmacogenomics. 2014;15(15):1867–79.

15. Wang JH. Effect of CYP3A4*1G, CYP3A5*3 genotype on serum carbamazepine concentrations at steady-state and carbamazepine efficacy in Han Chinese epileptic patients: FudanUniversity; 2012.

16. Ma CL, Jiao Z, Wu XY, et al. Association between PK/PD-involved gene polymorphisms and carbamazepine-individualized therapy. Pharmacogenomics. 2015;16(13):1499–512.

17. Yun W, Zhang F, Hu C, et al. Effects of EPHX1, SCN1A and CYP3A4 genetic polymorphisms on plasma carbamazepine concentrations and pharmacoresistance in Chinese patients with epilepsy. Epilepsy Res. 2013;107(3):231–7.

18. Liu X, Hu K, Gong L, et al. Effect of cytochrome P450 3A4 and 3A5 genotype on carbamazepine serum concentrations at steady-state and carbamazepine-incluced cutaneous adverse drug reactions. Chin J Clin Pharmacol. 2016;32(19):1749–52.

19. Seo T, Nakada N, Ueda N, et al. Effect of CYP3A5*3 on carbamazepine pharmacokinetics in Japanese patients with epilepsy. Clinical pharmacology and therapeutics. 2006;79(5):509–10.

20. Park PW, Seo YH, Ahn JY, Kim KA, Park JY. Effect of CYP3A5*3 genotype on serum carbamazepine concentrations at steady-state in Korean epileptic patients. Journal of clinical pharmacy and therapeutics. 2009;34(5):569–74.

21. Ambrósio AF, Soares-Da-Silva P, Carvalho CM, Carvalho AP. Mechanisms of action of carbamazepine and its derivatives, oxcarbazepine, BIA 2-093, and BIA 2-024. Neurochemical research. 2002;27(1-2):121–30.

22. Lasoń W, Chlebicka M, Rejdak K. Research advances in basic mechanisms of seizures and antiepileptic drug action. Pharmacological reports : PR. 2013;65(4):787–801.

23. Kwan P, Sills GJ, Brodie MJ. The mechanisms of action of commonly used antiepileptic drugs. Pharmacology & therapeutics. 2001;90(1):21–34.

24. Ebrahimi A, Houshmand M, Tonekaboni SH, et al. Two novel mutations in SCN1A gene in Iranian patients with epilepsy. Archives of medical research. 2010;41(3):207–14.

25. Meisler MH, O’Brien JE, Sharkey LM. Sodium channel gene family: epilepsy mutations, gene interactions and modifier effects. The Journal of physiology. 2010;588(Pt 11):1841–8.

26. Haerian BS, Baum L, Kwan P, et al. SCN1A, SCN2A and SCN3A gene polymorphisms and responsiveness to antiepileptic drugs: a multicenter cohort study and meta-analysis. Pharmacogenomics. 2013;14(10):1153–66.

27. Qian L, Fang S, Yan YL, et al. The ABCC2 c.-24C>T polymorphism increases the risk of resistance to antiepileptic drugs: A meta-analysis. Journal of clinical neuroscience : official journal of the Neurosurgical Society of Australasia. 2017;37:6–14.

28. Bao Y, Liu XZ, Xiao Z. Association between two SCN1A polymorphisms and resistance to sodium channel blocking AEDs: a meta-analysis. Neurological sciences : official journal of the Italian Neurological Society and of the Italian Society of Clinical Neurophysiology. 2018;39(6):1065–72.

29. Tate SK, Singh R, Hung C-C, et al. A common polymorphism in the SCN1A gene associates with phenytoin serum levels at maintenance dose. Pharmacogenet Genomics. 2006;16(10):721–6.

30. Zhou BT, Zhou QH, Yin JY, et al. Comprehensive analysis of the association of SCN1A gene polymorphisms with the retention rate of carbamazepine following monotherapy for new-onset focal seizures in the Chinese Han population. Clinical and experimental pharmacology & physiology. 2012;39(4):379–84.

31. Abe T, Seo T, Ishitsu T, et al. Association between SCN1A polymorphism and carbamazepine-resistant epilepsy. British journal of clinical pharmacology. 2008;66(2):304–7.

32. Liberati A, Altman DG, Tetzlaff J, et al. The PRISMA statement for reporting systematic reviews and meta-analyses of studies that evaluate healthcare interventions: explanation and elaboration. BMJ. 2009.

33. Kwan P, Arzimanoglou A, Berg AT, et al. Definition of drug resistant epilepsy: consensus proposal by the ad hoc Task Force of the ILAE Commission on Therapeutic Strategies. Epilepsia. 2010;51(6):1069–77.

34. Stang A. Critical evaluation of the Newcastle-Ottawa scale for the assessment of the quality of nonrandomized studies in meta-analyses. European journal of epidemiology. 2010;25(9):603–5.

35. Lin LF. Comparison of four heterogeneity measures for meta-analysis. Journal of evaluation in clinical practice. 2019:10.1111/jep.13159.

36. Meng HM, Ren JY, Lv YD, Lin WH, Guo YJ. Association study of CYP3A5 genetic polymorphism with serum concentrations of carbamazepine in Chinese epilepsy patients. Neurology Asia. 2011;16(1):39–45.

37. Liu JF, Li YC, Zhang GL, Tang Q. Effects of CYP3A5 Gene Polymorphism on Steady State Serum Concentrations and Therapeutic Efficacy of Carbamazepine in Chinese Han Epileptic Patients. China Pharmacy. 2014;25(36):3433–5.

38. Wang P, Zhou QH, Sheng YH, et al. Association between two functional SNPs of SCN1A gene and efficacy of carbamazepine monotherapy for focal seizures in Chinese Han epileptic patients. Zhong nan da xue xue bao. Yi xue ban = Journal of Central South University. Medical sciences. 2014;39(5):433–41.

39. Zhang Q, Tang H, Ji LH, Zhang HY, Li L. Effects of CYP3A5*3 Genetic Polymorphisms on Serum Concentrations in People with Epilepsy. Progress in Modern Biomedicine. 2016(27):5262–5.

40. Zhu YM, Hu JP, Zhu YL, Lu QS. Association of individual different plasma Carbamazepine and 10, 11-carbamazepine Epoxide concentration with CYP3A5*3 gene polymorphism. J Apoplexy and Nervous Diseases. 2012;29(9):804–7.

41. Daci A, Beretta G, Vllasaliu D, et al. Polymorphic Variants of SCN1A and EPHX1 Influence Plasma Carbamazepine Concentration, Metabolism and Pharmacoresistance in a Population of Kosovar Albanian Epileptic Patients. PLoS One. 2015;10(11):e0142408.

42. Hung CC, Chang WL, Ho JL, et al. Association of polymorphisms in EPHX1, UGT2B7, ABCB1, ABCC2, SCN1A and SCN2A genes with carbamazepine therapy optimization. Pharmacogenomics. 2012;13(2):159–69.

43. Sterjev Z, Kiteva G, Cvetkovska E, et al. Influence of the SCN1A IVS5N + 5 G>A Polymorphism on Therapy with Carbamazepine for Epilepsy. Balkan J Med Genet. 2012.

44. Huang JM, Qian Z, Chen HY, et al. Association of single nucleotide polymorphisms of SCNlA gene with therapeutic effect of carbamazepine among ethnic Zhuang Chinese patients with epilepsy. Chin J Med Genet. 2019;36(3):271–4.

45. Nazish HR, Ali N, Ullah S. The possible effect of SCN1A and SCN2A genetic variants on carbamazepine response among Khyber Pakhtunkhwa epileptic patients, Pakistan. Ther Clin Risk Manag. 2018.

46. Namazi S, Azarpira N, Javidnia K, et al. SCN1A and SCN1B gene polymorphisms and their association with plasma concentrations of carbamazepine and carbamazepine 10, 11 epoxide in Iranian epileptic patients. Iran J Basic Med Sci. 2015.

47. Cumpston M, Li T, Page MJ, et al. Updated guidance for trusted systematic reviews: a new edition of the Cochrane Handbook for Systematic Reviews of Interventions. The Cochrane database of systematic reviews. 2019;10:ED000142–ED.

48. Vesell ES. Genetic and environmental factors causing variation in drug response. Mutation research. 1991;247(2):241–57.

49. McLeod HL, Evans WE. Pharmacogenomics: unlocking the human genome for better drug therapy. Annual review of pharmacology and toxicology. 2001;41:101–21.

50. Mantegazza M, Curia G, Biagini G, Ragsdale DS, Avoli M. Voltage-gated sodium channels as therapeutic targets in epilepsy and other neurological disorders. The Lancet. Neurology. 2010;9(4):413–24.

51. Mulley JC, Scheffer IE, Petrou S, et al. SCN1A mutations and epilepsy. Human mutation. 2005;25(6):535–42.

52. Baulac S, Gourfinkel-An I, Picard F, et al. A second locus for familial generalized epilepsy with febrile seizures plus maps to chromosome 2q21-q33. American journal of human genetics. 1999;65(4):1078–85.

53. Moulard B, Guipponi M, Chaigne D, et al. Identification of a new locus for generalized epilepsy with febrile seizures plus (GEFS+) on chromosome 2q24-q33. American journal of human genetics. 1999;65(5):1396–400.

54. Copley J, Ziviani J. Barriers to the use of assistive technology for children with multiple disabilities. Occupational therapy international. 2004;11(4):229–43.

55. Tate SK, Depondt C, Sisodiya SM, et al. Genetic predictors of the maximum doses patients receive during clinical use of the anti-epileptic drugs carbamazepine and phenytoin. Proc Natl Acad Sci U S A. 2005;102(15):5507–12.

56. Thompson CH, Kahlig KM, George AL, Jr. SCN1A splice variants exhibit divergent sensitivity to commonly used antiepileptic drugs. Epilepsia. 2011;52(5):1000–9.

57. Sarao R, Gupta SK, Auld VJ, Dunn RJ. Developmentally regulated alternative RNA splicing of rat brain sodium channel mRNAs. Nucleic acids research. 1991;19(20):5673–9.

58. Gustafson TA, Clevinger EC, O’Neill TJ, Yarowsky PJ, Krueger BK. Mutually exclusive exon splicing of type III brain sodium channel alpha subunit RNA generates developmentally regulated isoforms in rat brain. The Journal of biological chemistry. 1993;268(25):18648–53.

59. Tan J, Liu Z, Nomura Y, Goldin AL, Dong K. Alternative splicing of an insect sodium channel gene generates pharmacologically distinct sodium channels. The Journal of neuroscience : the official journal of the Society for Neuroscience. 2002;22(13):5300–9.

60. Fletcher EV, Kullmann DM, Schorge S. Alternative splicing modulates inactivation of type 1 voltage-gated sodium channels by toggling an amino acid in the first S3-S4 linker. The Journal of biological chemistry. 2011;286(42):36700–8.

61. Yip TSC, O’Doherty C, Tan NCK, Dibbens LM, Suppiah V. SCN1A variations and response to multiple antiepileptic drugs. The pharmacogenomics journal. 2014;14(4):385–9.

62. Lakhan R, Kumari R, Misra UK, et al. Differential role of sodium channels SCN1A and SCN2A gene polymorphisms with epilepsy and multiple drug resistance in the north Indian population. British journal of clinical pharmacology. 2009;68(2):214–20.

63. Abo El Fotoh WMM, Abd El Naby SAA, Habib MSE-D, Alrefai AA, Kasemy ZA. The potential implication of SCN1A and CYP3A5 genetic variants on antiepileptic drug resistance among Egyptian epileptic children. Seizure. 2016;41:75–80.

64. Caruso A, Bellia C, Pivetti A, et al. Effects of EPHX1 and CYP3A4 polymorphisms on carbamazepine metabolism in epileptic patients. Pharmgenomics Pers Med. 2014;7:117–20.

65. Rogers JF, Nafziger AN, Bertino JS, Jr. Pharmacogenetics affects dosing, efficacy, and toxicity of cytochrome P450-metabolized drugs. The American journal of medicine. 2002;113(9):746–50.

66. Glue P, Clement RP. Cytochrome P450 enzymes and drug metabolism--basic concepts and methods of assessment. Cellular and molecular neurobiology. 1999;19(3):309–23.

67. Huang W, Lin YS, McConn DJ, 2nd, et al. Evidence of significant contribution from CYP3A5 to hepatic drug metabolism. Drug Metab Dispos. 2004;32(12):1434–45.

68. He BX, Shi L, Qiu J, et al. A functional polymorphism in the CYP3A4 gene is associated with increased risk of coronary heart disease in the Chinese Han population. Basic & clinical pharmacology & toxicology. 2011;108(3):208–13.

69. Ozeki T, Nagahama M, Fujita K, et al. Influence of CYP3A4/5 and ABC transporter polymorphisms on lenvatinib plasma trough concentrations in Japanese patients with thyroid cancer. Scientific reports. 2019;9(1):5404-.

70. Hustert E, Haberl M, Burk O, et al. The genetic determinants of the CYP3A5 polymorphism. Pharmacogenetics. 2001;11(9):773–9.

71. Li CJ, Li L, Lin L, et al. Impact of the CYP3A5, CYP3A4, COMT, IL-10 and POR genetic polymorphisms on tacrolimus metabolism in Chinese renal transplant recipients. PloS one. 2014;9(1):e86206–e.

72. Ghosh C, Gonzalez-Martinez J, Hossain M, et al. Pattern of P450 expression at the human blood-brain barrier: roles of epileptic condition and laminar flow. Epilepsia. 2010;51(8):1408–17.

73. Kuehl P, Zhang J, Lin Y, et al. Sequence diversity in CYP3A promoters and characterization of the genetic basis of polymorphic CYP3A5 expression. Nature genetics. 2001;27(4):383–91.

